# Tradeoff between speed and reproductive number in pathogen evolution

**DOI:** 10.1101/2022.06.30.22277094

**Authors:** Andreas Eilersen, Bjarke Frost Nielsen, Kim Sneppen

## Abstract

The rapid succession of new variants of SARS-CoV-2 emphasizes the need to understand the factors driving pathogen evolution. Here, we investigate a possible tradeoff between the rate of progression of a disease and its reproductive number. Using an SEIR framework, we show that in the exponential growth phase of an epidemic, there is an optimal disease duration that balances the advantage of a fast disease progression with that of causing many secondary infections. This result offers one possible explanation for the ever shorter generation times of novel variants of SARS-CoV-2, as it progressed from the original strain to the Alpha, Delta, and, from late 2021 onwards, to several Omicron variant subtypes. In the endemic state, the optimum disappears and longer disease duration becomes advantageous for the pathogen. However, selection pressures depend on context: mitigation strategies such as quarantine of infected individuals may slow down the evolution towards longer-lasting, more infectious variants. This work then suggests that, in the future, the trend towards shorter generation times may reverse, and SARS-CoV-2 may instead evolve towards longer-lasting variants.

## INTRODUCTION

Since the emergence of SARS-CoV-2, multiple variants of the virus with faster transmission dynamics have arisen. The variants have supplanted each other in successive waves, with variants with ever higher transmission rates and/or shorter generation times replacing older, slower variants [1, 2]. This unfolding evolutionary race suggests a dynamic that can be explored through modeling. Here, we explore the tradeoff between the number of secondary cases an epidemic disease has time to cause over its infectious period and the speed with which the pathogen goes through disease generations.

Some work has already been done on modeling the evolution of the infection profile of SARS-CoV-2 and other pathogens with similar generation times. Saad-Roy *et al*. studied the evolution of a presymptomatic infectious state under the assumption that such a state is less infectious [3], and in the context of superinfection and within-host competition [4]. In addition, the relationship between the duration of a disease or parasitic infection and the infection rate has been studied under the assumption of a tradeoff between the two given by some functional relationship [5, 6]. Porco *et al*. [7] investigated the effects of treatment and other interventions on disease evolution under the assumption of a similar tradeoff. Analogous studies have been done on other ecological relationships, such as predation [8, 9]. Finally, Park *et al*. [10] have studied the interplay between disease infectivity and speed with a focus on mitigation rather than evolution. However, the possibility that a longer infectious period might be an evolutionary advantage for a disease only up to a certain threshold has not been studied in detail.

Here we investigate disease duration alone, and not increases in the infection rate, which confer an obvious advantage for the pathogen, while the situation is less obvious when it comes to the rate of disease progression. We focus on the tradeoff between the duration of the individual infections and the number of secondary cases that each infected individual generates. We assume that infected individuals transmit the disease at a constant rate during the infectious period. This means that a long disease duration should lead to a higher effective reproductive number, *R*_0_, that is, to more secondary infections. On the other hand, a long disease duration might also be a disadvantage to the disease, as it may be associated with a long latency and thereby a slow epidemic progression. This is particularly the case if one assumes direct proportionality between the duration of the latency time of a disease and its infectious period. Table 1 suggests that across diseases spread through the air or via direct social contact, longer infectious periods indeed correlate with longer latent periods. In a susceptible-exposed-infectious-recovered (SEIR) type compartmental model, which we will consider in this study, the latent period corresponds to the *E*-state. We will derive relations and carry out epidemic simulations based on systems of ordinary differential equations (ODEs) to investigate the condition for an optimum disease duration.

**TABLE 1.**
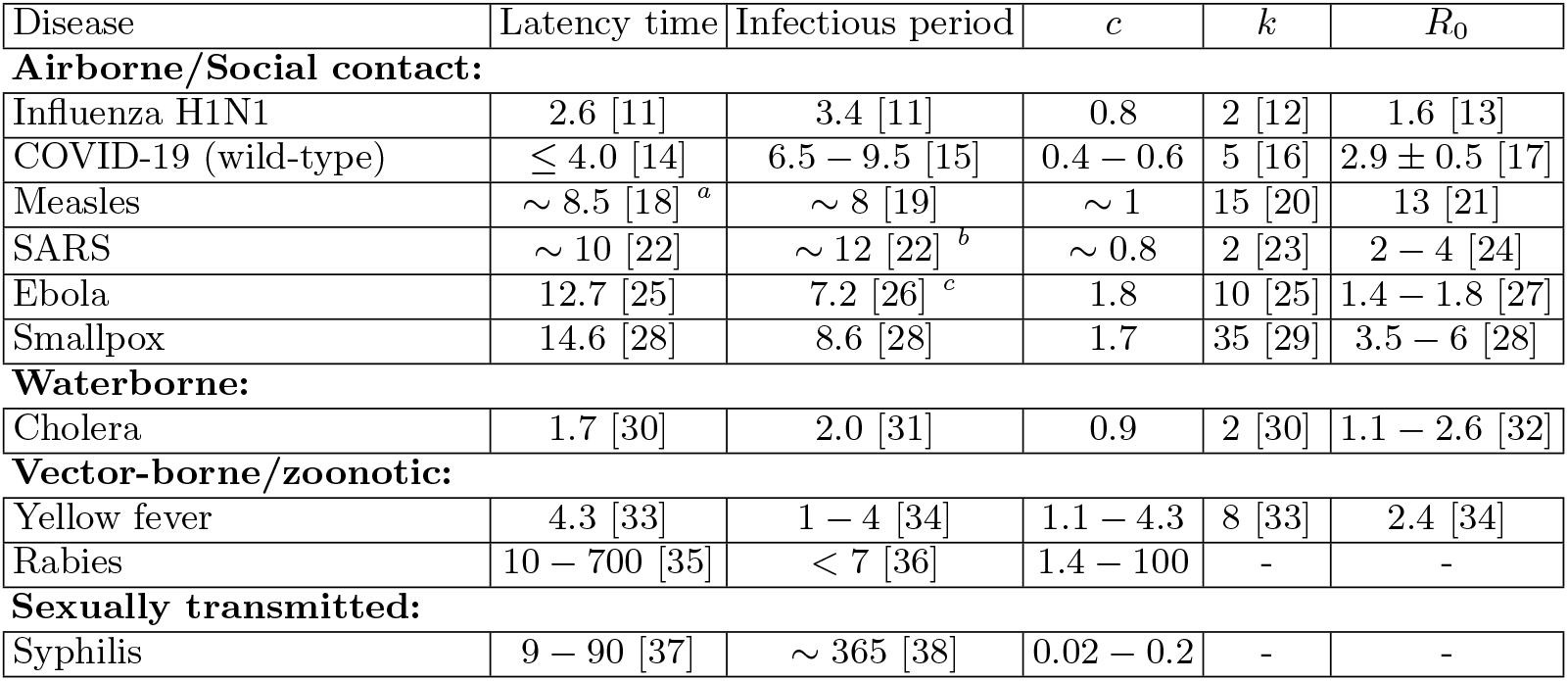
Duration (days) of the latent and infectious periods for some infectious diseases. *c* is the ratio of the latency time to the infectious period of the disease, while *k* = *k*_*incubation*_ is the shape factor for a Gamma distribution which we fit approximately to the measured distribution of incubation times found in the cited literature. The coefficient of variation of these distributions is given by *CV*^2^ = 1*/k*. Note that we show the *k*-values for the incubation periods as opposed to the latent periods, which we assume are similarly distributed. The incubation period is the time from infection to symptom onset, while the latent period is the time from infection to onset of infectiousness. The last column shows the estimated basic reproductive number. Diseases are here sorted by whether they are mainly spread through water, by vectors or other animals, by sexual contact, or by droplets or aerosols upon casual social contact. ^*a*^ As infectiousness begins four days before the onset of a rash, latency time is calculated as incubation period minus four days. ^*b*^ Incubation period is reported as 4.6 days. Here, we define the infectious period by the requirement that at least 50 % of patients secrete measurable quantities of the virus. In that case, infectiousness begins on day five after the onset of symptoms and ends on day 17, yielding a latency time of 9.6 days. ^*c*^ The cited study reports separate infectious periods for survivors and deceased patients. We have here indicated the average.

## MODEL SETUP

We assume that the transmission rate *β* of a disease is constant throughout the duration of the infectious period *T*, giving a linear relationship between disease duration and number of secondary cases. While strict proportionality does not necessarily hold, [39, 40] a positive, monotonic relation between the two is expected since a longer infectious period leads to more opportunities for passing on the infection.

Throughout this work, we will distinguish between the latent and incubation periods of a disease. The latent period is the time from the initial infection until the patient becomes infectious. In contrast, the incubation period is the time from infection until the onset of symptoms.

The SEIR model reads:

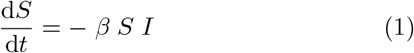

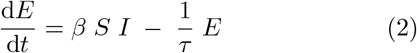

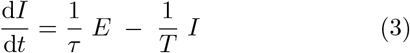

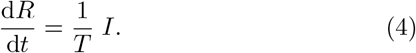

where *S, E, I*, and *R* are susceptible, exposed (but non-infectious), infectious, and recovered compartments respectively. *β* is the transmission rate per unit time, *τ* is the average duration of the pre-infectious exposed period, and *T* is the duration of the infectious period. We take the total population of the system to be fixed at *N* = 1 and assume that the time-scale of the entire scenario is short enough that vital dynamics - births and deaths - can be neglected.

We will also investigate the effects of variability in the durations of the *E* and *I* states. Usually, an exponential distribution is assumed in SEIR models. However, by subdividing each of the compartments *E* and *I* into *k* equally long sub-compartments, we instead obtain a Gamma distribution of latency times and infectious periods with a shape parameter *k* [41]. The shape parameter is related to the coefficient of variation *CV* by 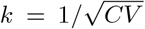. Thus, a greater *k* corresponds to lower person-to-person variation in the durations of the *E* and *I* states. Since we assume integer values of *k*, this distribution coincides with an Erlang distribution. The above equations for the simple SEIR model correspond to letting the shape parameter take on the value *k* = 1, yielding the familiar exponential case.

When considering the evolution of the disease in an endemic state with a high degree of existing immunity in the population, we slightly modify the above SEIR model. To make the endemic state possible, we allow individuals to lose immunity at a rate *ω*, corresponding to an SEIRS model of disease progression. This model is then solved numerically, including multiple co-circulating variants with different disease durations *T*. Thereby we emulate the natural competition between variants.

Finally, mitigation by isolation of infected individuals is implemented by the addition of a quarantine rate *q* and corresponding noninfectious quarantine compartment *Q* to the SEIRS model. This represents how individuals have some chance of becoming symptomatic, being contact traced, or otherwise being diagnosed and isolated for each day of illness. We expect that individuals suffering from a very long-lasting infectious disease will eventually self-quarantine. The resulting equations thus become

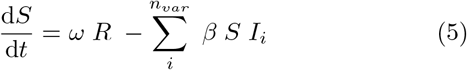

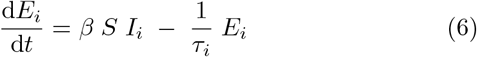

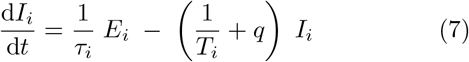

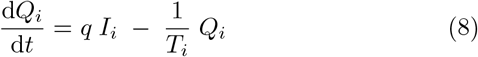

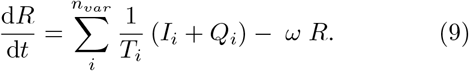

Here we have introduced an index *i* to indicate the possibility of including *n*_*var*_ different variants, all of which we assume to have perfect cross immunity with respect to the others. This will become important when simulating competition between strains with different disease durations and latency times.

## RESULTS

### Optimum disease duration for exponential growth

In the exponential growth phase of an epidemic, the growth rate for *k* = 1 may be determined by linearizing the system of equations (1-4) around the disease-free equilibrium. The epidemic growth rate *r* is then the largest eigenvalue of the Jacobian [42]:

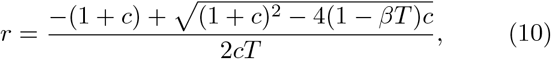

where *c* ≡ *τ/T*, and we have chosen the physically allowable positive branch. The above function has a maximum for the duration *T* given by

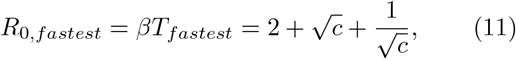

In the limit of *k* → ∞, the durations of the exposed and infectious stages are deterministic. If we further neglect the variation in the timing of disease transmission, total disease duration thus becomes *τ* +*T* and the generation time becomes *τ* + *T/*2. Under these approximations, the number of infected in the exponential growth phase approaches

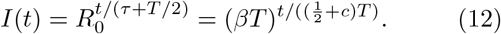

This is only an approximate expression, as it requires the assumption that disease transmission is also deterministic, which is not the case even for *k* → ∞. Maximising the exponential growth rate as a function of *T* in the infinite-*k* limit and under the assumptions of Eq. (12), we obtain the finite value

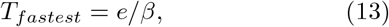

where *e* denotes Euler’s number.

In the exponential growth phase, the variant with the highest growth rate will quickly come to dominate. An illustration of this phase for different disease durations and *k* = *c* = 1 can be seen in Fig. 1, while a plot of the growth rate as a function of *T* for various values of *c* is shown in Fig. 2(a). As shown, the exponential growth rate is much higher for lower values of *c*. Furthermore, there is good agreement between our analytical and numerical calculations. When increasing *c*, the maximum growth rate decreases strongly, and the optimum with respect to *T* becomes less clear.

**FIG. 1.**
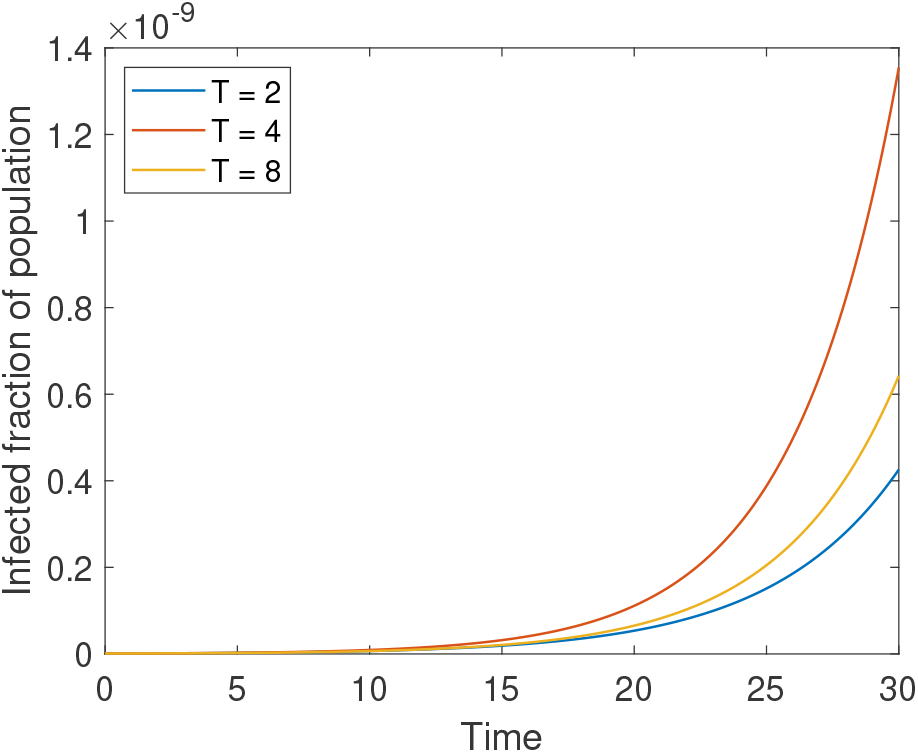
Direct simulation of the initial growth of an epidemic for different disease durations *T*. When disease duration is short, the epidemic grows faster as *T* increases because each infected individual can infect more people. However, for larger values of *T*, the cost of the longer latent period increases and leads to slower exponential growth. Here, *β* = 1, *k* = 1, and *c* = *τ/T* = 1, corresponding to assuming equal duration of the *E* and *I* states. The initial exposed and infected fractions of the population are *E*(0) = 10^−12^ and *I*(0) = 10^−12^.

**FIG. 2.**
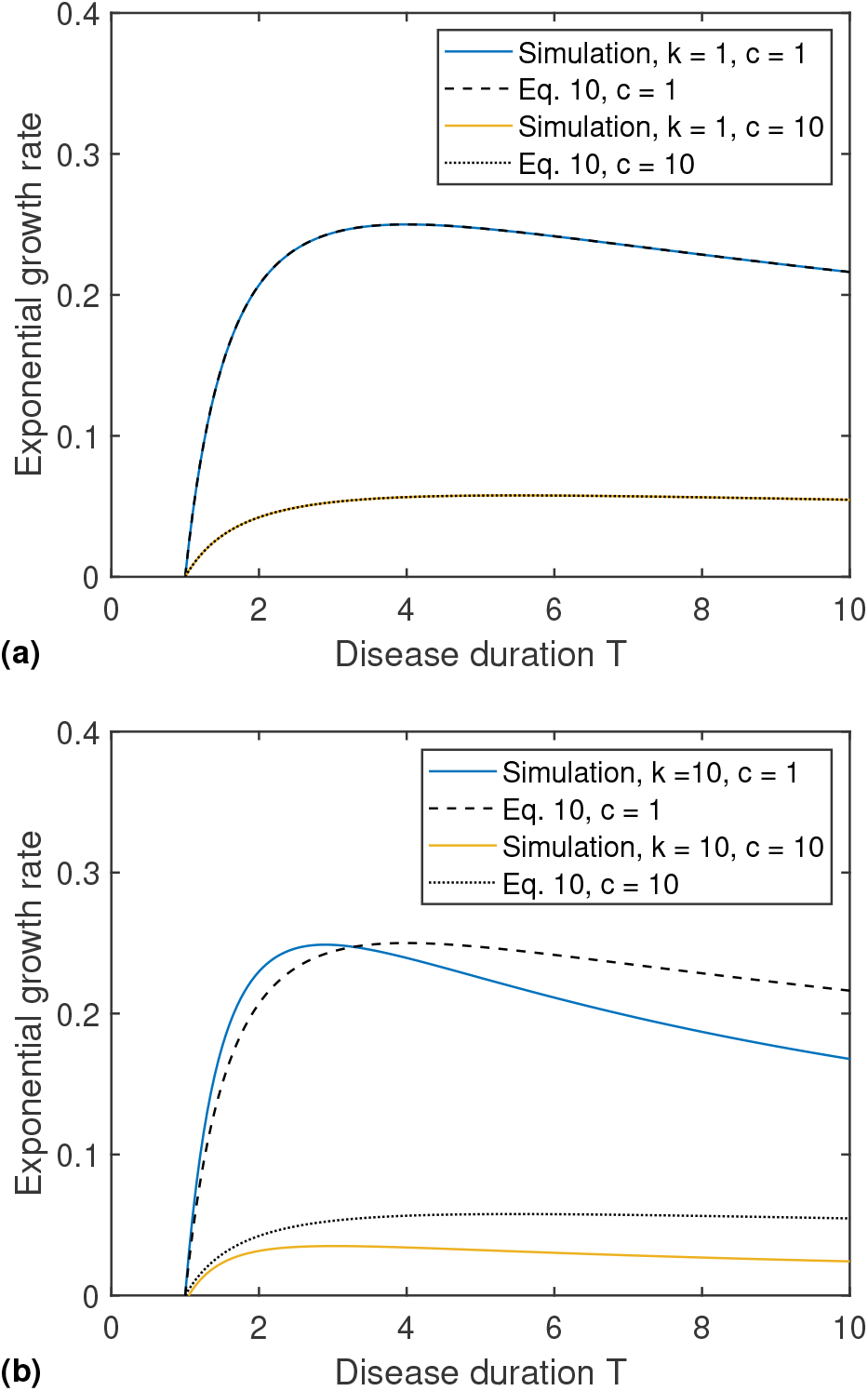
Growth rate as a function of infectious period duration *T* as simulated, compared with the growth rate derived in Eq. (10). Here, we set the infection rate *β* = 1*/*day. In panel (a) we vary the ratio between exposed and infectious period duration, *c*. One observes that the growth rate has a maximum at the duration given by Eq. (11) (giving *T* = 4 for *c* = 1 and *T* = 5.48 for *c* = 10). Panel (b) shows the corresponding plot when considering a larger shape factor *k* for the distribution of the durations of latent and infectious periods.

In the equation for *r* (Eq. (10)), it is assumed that the probability distribution for the duration of the latency time and infectious period of each individual is an exponential distribution, corresponding to a shape parameter *k* = 1. We wish to explore how the degree of variability in the latent and infectious period affects the optimal value disease duration and the resulting growth rate. We do this numerically by solving the SEIR equations with a Gamma distributed latent and infectious period and varying the shape parameter.

In Fig. 2(b), the effects of varying *k* are shown. At *c* = 1, the effect of an increase in *k* is modest. However, when the latent period is much longer than the infectious period (*c* = 10), a more definite duration (*k* = 10) is associated with a clearly lower resultant growth rate. A more complete overview of growth rates and maxima as functions of *c* for different *k* is given in Fig. 3.

**FIG. 3.**
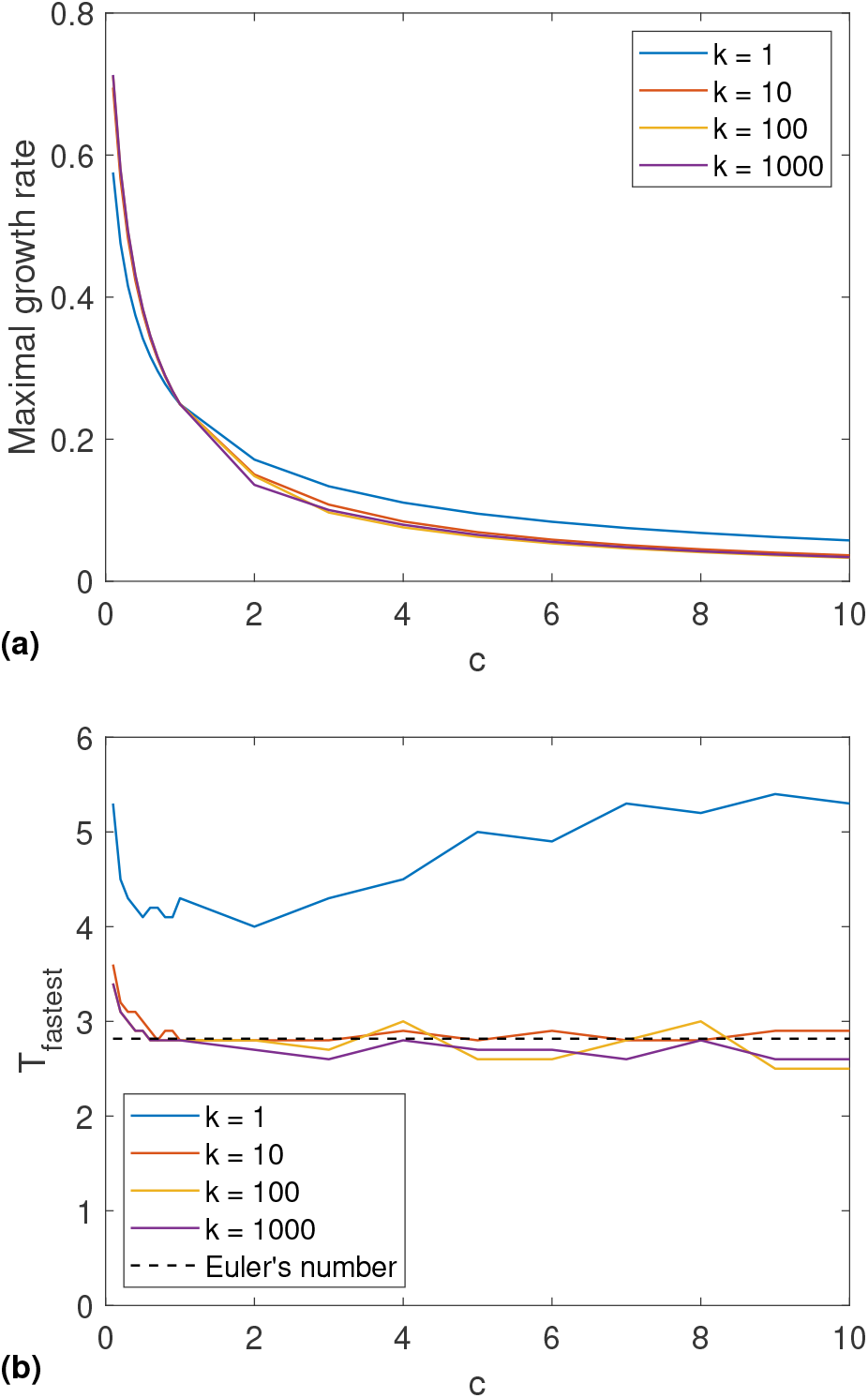
Maximal growth rates and optimal disease durations as functions of *c* for various *k*. (a) shows the maximal exponential growth rate *r* of an epidemic pathogen given different values of *c* and *k*. We see that growth rates are highest for low *c* and also decrease very slightly with *k*. (b) shows the disease duration *T*_*fastest*_ that maximizes the growth rate. For *β* = 1 this is also the value of *R*0 that maximizes the growth rate. This is equal to the predictions from Eq. (10) for *k* = 1 and drops as *k* increases. At higher *k, T*_*fastest*_ approaches *e* as predicted in the limit of *k* → ∞.

Overall, Fig. 3 illustrates that the maximal daily growth rate decreases monotonically as a function of *c*, reflecting a longer latency time. Increasing *k*, i.e., making the distribution of latency times more sharply peaked, also leads to a slight decrease in maximal growth rate for *c >* 1.

Fig. 3 (b) shows that the value of *T* which maximises growth rate *r*, denoted *T*_*fastest*_, is highest in the limit of *c* → 0. At low values of *k, T*_*fastest*_ exhibits a dependence on *c*, with a local minimum around *c* = 1 beyond which it grows as *c* increases. *T*_*fastest*_(*c*) for *k* = 1 reproduces Eq. (11). As *k* is increased, *T*_*fastest*_ becomes nearly independent of *c* and approaches a value of approximately *e*, as predicted by Eq. (13) for *β* = 1. The good agreement is remarkable given the simplifications contained in that equation.

The exponential growth rates of Fig. 3 are calculated by fitting an exponential function to the initial phase of a simulated SEIR epidemic model. For high values of *k* and *c*, the disease prevalence oscillates around the expected exponential curve over the course of a disease generation. In this case, we fit an exponential function to the local peaks rather than the whole curve. Due to these inherent fluctuations in the number of infected, the numerically determined exponential growth rate is very sensitive to the exact start- and end points of the fit. This gives rise to the slight fluctuations seen in the curves of 3(b).

In a situation where the disease is growing exponentially, e.g., when an epidemic is breaking out or control measures are failing, the variant with the fastest exponential growth rate will win as illustrated in Fig. 1. This, however, only holds transiently and we will see that the situation is reversed when considering the endemic state in a model with immunity loss.

### The endemic state

In the endemic state, the disease prevalence is sustained at an approximately constant level. In our model, this is done by introducing a small rate of loss from the recovered (*R*) state, corresponding to the waning of acquired immunity (terms involving *ωR* in Eqs. (5) and (9)). Furthermore, we will consider a quarantine rate *q* that quantifies the probability per day that an individual in the *I* state goes into isolation (see Eq. (7)). As previously described, this is represented by a noninfectious quarantine compartment *Q* which infected persons may leave upon recovery (see Eq. (8)). The extended multistrain model (Eqs. (5)-(reduces to the simple SEIR model of Eqs. (1-4) when only one strain is included and *q* = *ω* = 0. If one were to consider very long disease durations, inclusion of vital dynamics (birth and death) would be necessary. This would entail the inclusion of loss terms from the *E, I*, and *Q* states. This would in turn limit the maximal effective duration of diseases to be below the scale of a human generation.

The results of the simulations including immunity loss and quarantine are illustrated in Fig. 4. The figure shows that in this case, longer-lasting variants always outcompete shorter-lasting ones. The simulations illustrate that this pattern persists even for very long disease durations (up to 400 days), although the replacement dynamics become extremely slow when, e.g., a 300-day and a 400-day variant compete (simulation time *>* 10^5^days). If the system starts out with an equal fraction of the population infected with each variant, we thus see a succession of variant takeovers (Fig. 4(a)). Longer-lasting variants take longer to grow, but eventually always end up taking over due to their higher basic reproductive number. However, by increasing the quarantine rate *q* (Fig. 4(b)), we see that this development can be slowed significantly. We have also examined the sensitivity to variations in the immunity loss rate *ω*, but varying this rate has little effect, except on the magnitude of outbreaks.

**FIG. 4.**
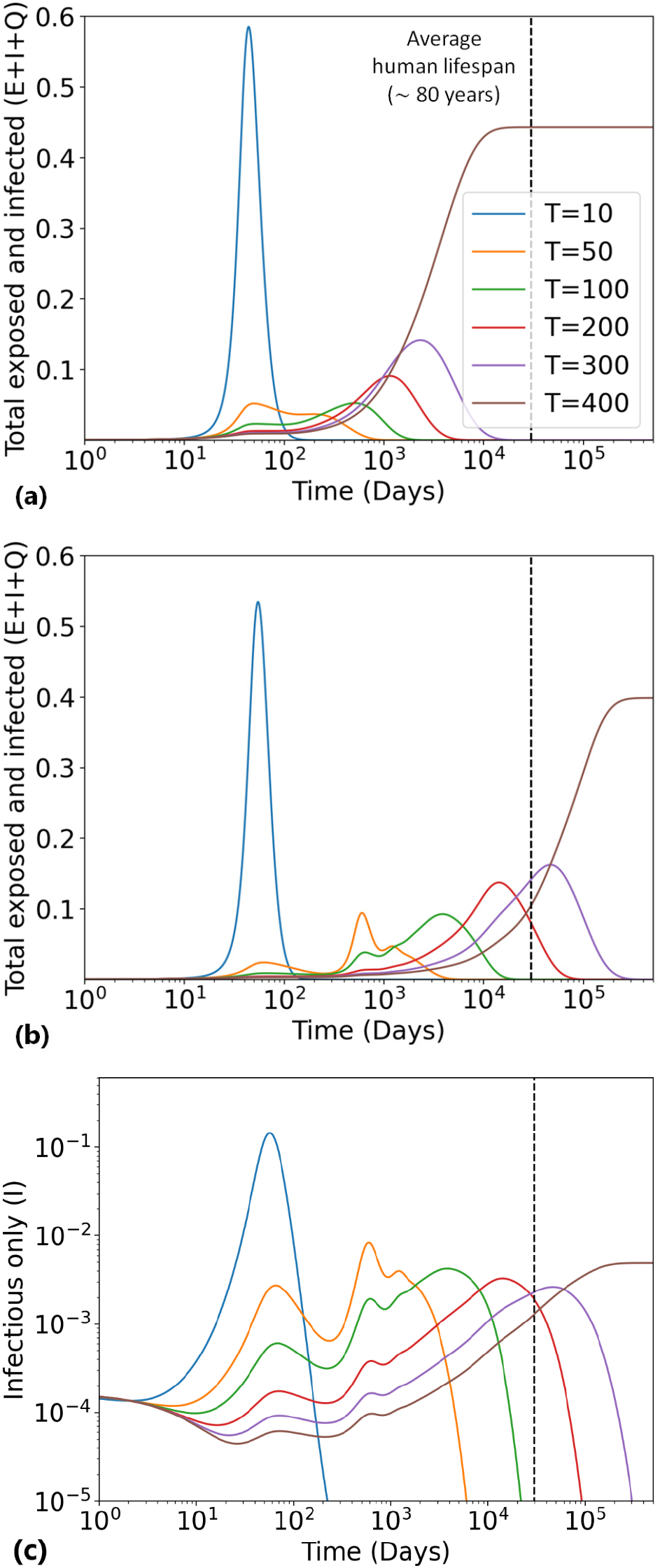
Competition between variants of different infectious period duration *T* as the disease progresses to the endemic phase. We here show the results of an ODE-based simulation of six different variants of duration *T* = 10, 50, 100, 200, 300, 400 days. The simulations use fixed *β* = 1, *c* = 1, and *ω* = 10^−3^. The first two panels show the total exposed, infected, and quarantined infected population *E* + *I* + *Q*. In (a), we simulate the system without quarantine. We see that the longer-lasting variants replace the shorter-lived ones. (b) shows a simulation using the same parameters and a quarantine rate of *q* = 0.1 per day of infectious illness. The longer-lasting variants still win, but quarantine slows down the evolution. (c) shows the same simulation as (b), but only *I* is plotted. *I* decreases as longer-lasting variants take over, since more and more of the population is quarantined for very long disease durations.

In the limit of very large *T* (formally, *T* → ∞), the size of the recovered population *R* goes to 0, while the susceptible population tends towards the limit *S* → *q/β*. Thus *E* + *Q* + *I* → 1 − *q/β*. Interestingly, in this limit the vast majority of non-susceptible individuals will be in the exposed and quarantined states, since, at steady state *I/*(*E* + *Q*) = 1*/*(*c* + (1 + *c*)*Tq*) which decreases as *T* is increased. Strictly speaking, the *T* → ∞ limit is only meaningful if vital dynamics are included, but already at moderate values of *T*, we can see that *E* + *Q* + *I* increases with increasing *T* while *I* in fact decreases (see Fig. 4(c)). As such, the competitive advantage of slower variants does not owe to a higher number of individuals in the *I* state, but rather to a minimization of the uninfected population.

## DISCUSSION

Our analysis illustrates that being fast-acting can be an evolutionary advantage for a pathogen, even if it comes at the cost of a lower reproductive number. However, this is only the case in the initial exponential growth phase of the epidemic. In the endemic phase the longer-lasting variants which have a higher *R*_0_ will always eventually come to dominate. This is the case regardless of interventions such as quarantine, though evolution towards long-lasting, more infectious variants may be slowed by quarantining infectious individuals.

We expect the two scenarios modelled here to be applicable across most of the trajectory of a real-world epidemic. With regard to quarantine, we would expect the onset of symptoms to increase the chance that individuals stay home or are bedridden, effectively self-quarantining. This is likely the case regardless of large-scale mitigation policies.

In addition, the exponential growth scenario is not necessarily limited to the short initial stage of the epidemic. In the case of the COVID-19 pandemic, mitigation efforts in various locations often kept the local reproductive number at or below 1. When such efforts failed or were relaxed, local epidemics entered a new exponential growth phase. Our results may thus contribute to an understanding of the successive shifts from the Wuhan strain, to the Alpha and Delta variants, and then, during 2022, to various Omicron subtypes. The Delta variant has been shown to have a somewhat shorter incubation period and significantly shorter generation time than the ancestral strain [43–45] Hart *et al*. [46] measure a generation time of 5.5 days for the Alpha variant and 4.6 days for the Delta variant. Omicron was even faster, with a reported serial interval of only 2.2 days [47]. The analysis in Abbott *et al*. further supports the tendency of faster disease progression for the latter SARS-CoV-2 variant, although Pung *et al*. dispute whether generation times of Delta were in fact significantly lower than for the Alpha variant [2, 48].

Each new SARS-CoV-2 variant has been accompanied by changes in transmission rate as well as generation time. The analysis presented here focuses on the time aspect while ignoring the obvious evolutionary gain a pathogen may obtain by increasing the infection rate *β*. We found that the growth rate in the exponential growth phase of an epidemic indeed seems to be optimal for rather short generation times, which agrees with the obervation that new SARS-CoV-2 variants tend to be faster than older variants. This overall tendency during 2020-2021 with ever faster virus variants may later be broken, however. Our simulations demonstrate that this is likely if the pandemic reaches a more endemic state where slower variants of the disease gain in fitness, as mitigation and quarantine efforts are dropped. Even under the assumption of quarantine measures, the optimal strategy should shift towards a longer disease duration in the endemic state, albeit more slowly.

Our analysis focuses on pathogens like SARS-CoV-2, which are transmitted through social contact and act on a relatively short timescale. There are of course also pathogens that act on timescales longer than those predicted here. This is for example true of sexually transmitted infections such as syphilis and HIV which cause lifelong infection (*c <<* 1). These infections violate the assumption of latency time being roughly proportional to infectious period duration, and thus are not captured by our model. In the case of these STIs, the two aspects of pathogen dynamics are essentially decoupled in the body.

Alternative approaches have been used to investigate the reasons for the variation in disease duration observed in the real world. These have predicted the existence of several “regimes” of disease duration [49]: from fast-acting childhood infections in situations with high contact rates to lifelong infections in low-contact situations.

It is of course both idealized and highly simplified to assume that latency times scale proportionally with infectious periods. As mentioned in the introduction, there is however some support for our assumption of a relationship between the two quantities. The values of *c* are indeed often on the order of 1, except for zoonotic diseases like rabies or sexually transmitted infections like syphilis (Table 1). Interestingly, the data show a large variation in how sharply peaked the incubation periods are, with the shape parameter *k* varying from ≈ 2 in respiratory diseases such as SARS and influenza, to an estimated 35 in smallpox.

The findings of this article highlight the context-dependence of evolutionary fitness as it pertains to disease duration and latency. Our results may help explain some of observed dynamics of emerging SARS-CoV-2 variants. In a wider perspective, our work also sheds some light on the apparent division of infectious diseases into a group of quite fast diseases characterized by epidemic outbreaks, and another group which are slow with long latent periods and an endemic pattern of infection.

## Data Availability

The code used to generate the plots shown in
this article is available on GitHub under the URL
https://github.com/gks973/Evolution_of_speed
or in a permanent archived version at
https://doi.org/10.5281/zenodo.7512664.

## ACKNOWLEDGMENTS

We wish to thank Lone Simonsen, Viggo Andreasen, and Nils Christian Stenseth for enlightening discussions.

KS and AE have received funding from the European Research Council (ERC) under the European Union’s Horizon 2020 research and innovation programme (grant agreement no. 740704). BFN received funding from the Carlsberg Foundation under its Semper Ardens programme (grant no. CF20-0046), and AE received funding from NordForsk under the Nordic Programme for Interdisciplinary Research (grant no. 104910).

## DATA AVAILABILITY

The code used to generate the plots shown in this article is available on GitHub under the URL https://github.com/gks973/Evolution_of_speed or in a permanent archived version at https://doi.org/10.5281/zenodo.7512664.

## AUTHOR CONTRIBUTIONS STATEMENT

KS proposed the model. AE, KS and BFN derived the expressions, and BFN and AE wrote the code for simulations. KS, AE, and BFN wrote the manuscript.

